# Development of a Monoclonal Antibody and a Sandwich-ELISA for the Detection of Mucormycosis in Humans

**DOI:** 10.64898/2026.04.23.26351301

**Authors:** Christopher R. Thornton, Genna E. Davies

## Abstract

**Background:** Mucormycosis is a rapidly progressive and often fatal invasive fungal infection caused by moulds in the order, *Mucorales*. Early diagnosis is essential for effective clinical management; however, conventional diagnostic approaches such as culture and histopathology are slow, insensitive, and require specialist mycological expertise. Although molecular methods are available for disease detection, they are not widely accessible. At present, no enzyme immunoassay (EIA) exists for the detection of mucormycosis.

**Methods:** A murine IgG1 monoclonal antibody (mAb), FH12, was generated against extracellular polysaccharides (EPSs) produced by *Mucorales* pathogens during active growth. The antibody was characterised for specificity, epitope stability, and antigen localisation using ELISA, immunoblotting, and immunofluorescence techniques. The mAb was incorporated into a Sandwich-ELISA and evaluated using culture filtrates, purified EPSs spiked into human serum, and tissue homogenates from a patient with cutaneous mucormycosis caused by *Lichtheimia ramosa*.

**Results:** mAb FH12 demonstrated pan-*Mucorales* specificity and no cross-reactivity with other clinically relevant yeasts and moulds. The epitope recognised by FH12 is periodate-insensitive and moderately heat-stable. The Sandwich-ELISA detected EPS antigens in human serum with limits of detection ranging from pg/mL to low ng/mL levels, and successfully identified the EPS biomarker in patient tissue homogenates.

**Conclusion:** The FH12-based Sandwich-ELISA shows high sensitivity and specificity, and has the potential to be used as a laboratory-based adjunct diagnostic test for the detection of mucormycosis in humans.

## 1. Introduction

Mucormycosis is a rapidly-progressive angio-invasive disease of humans caused by fungi in the Zygomycete order, *Mucorales*, manifesting as life-threatening rhino-orbital-cerebral, pulmonary, cutaneous, gastrointestinal, and disseminated infections (Figure 1A) [1]. The species most commonly reported as pathogens of humans are *Apophysomyces variabilis, Cunninghamella bertholletiae, Lichtheimia corymbifera* and *Lichtheimia ramosa, Mucor circinelloides, Rhizomucor pusillus, Rhizopus arrhizus*, and *Rhizopus microsporus* [2]. Identified as a global high-priority pathogen group by the World Health Organisation [3], and a critical-priority pathogen group in South-East Asia by others [4], mortality rates from *Mucorales* infections of up to 80% have been reported [2].

**Figure 1.**
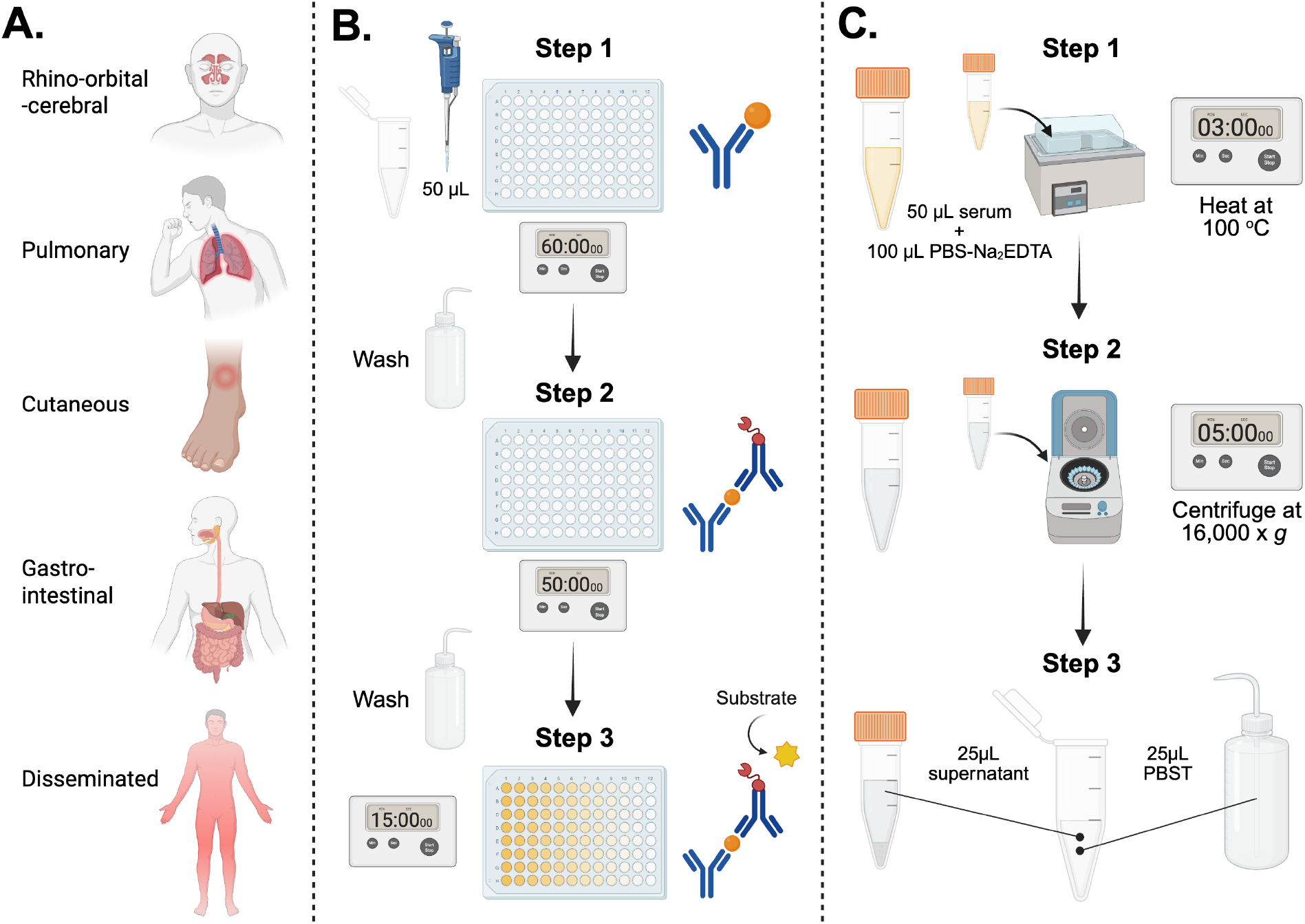
(**A**). Diseases caused by *Mucorales* fungi. The predominant disease in humans is rhino-orbital-cerebral mucormycosis (ROCM) involving the nose, eye and brain, and is typically reported in the settings of poorly controlled diabetes and haematological malignancy. A surge of ROCM was reported during the second wave of the Coronavirus pandemic, especially in diabetic patients and/or those receiving corticosteroids for COVID-19 infection. Pulmonary mucormycosis is most commonly seen in haematological malignancy and transplant patients, and is related to desferrioxamine therapy with renal impairment. Cutaneous (skin and soft tissue) mucormycosis can occur in burn wounds, and following contaminated traumatic injury, including tornado-type and combat-related injuries. Less common manifestations include gastro-intestinal and disseminated mucormycosis. (**B**). Sandwich-ELISA protocol. Step 1: Wells of microtiter plates coated with 50 μL of Protein A-purified mAb FH12 (4 μg/mL) are blocked for 15 min with 100 μL of PBS containing 1.0% (wt:vol) BSA, and then incubated at 23 °C for 1 h with 50 µL of antigen solution. Step 2: After washing, the wells are incubated for 50 min at 23 °C with FH12-HRP conjugate diluted 1 in 10,000 (vol:vol) in PBST containing 0.25% (wt:vol) BSA. Step 3: After washing, the wells are incubated with substrate solution for 15 min and the absorbance values determined at 450 nm. (**C**). Procedure for heat treatment of human serum. Step 1: human serum is mixed 1:2 (vol:vol) with PBS buffer containing Na^2^EDTA, and then heated for 3 min at 100 °C in a boiling water bath. Step 2: the heated serum is centrifuged at 16,000 x *g* for 5 min to pellet insoluble serum proteins. Step 3: following centrifugation, the clear supernatant is mixed 1:1 (vol:vol) with running buffer (RB) and 50 μL assayed by Sandwich-ELISA. Figure 1 was created with BioRender.com.

The prompt detection and differentiation of *Mucorales* species from other mould pathogens such as *Aspergillus fumigatus* is critical to patient survival, enabling timely treatment with surgery and with *Mucorales*-active antifungal drugs [5]. Detection of *Mucorales* pathogens has traditionally relied on histopathology and fungal culture from a biopsy sample, and the identification by microscopy of characteristic morphological structures, but these procedures are laborious, insensitive and require mycological expertise [5,6]. For this reason, more sophisticated and costly diagnostic modalities such as metagenomic next-generation sequencing (NGS) [7] or matrix assisted laser desorption ionization time of flight (MALDI-TOF) [8] can be used for their identification following culture *in vitro*, but access to such technology is limited.

While polymerase chain reaction (PCR) tests have been developed for the detection of circulating *Mucorales* nucleic acid in human serum and bronchoalveolar lavage fluid [9,10], there is currently no enzyme immunoassay (EIA) that can be used as adjunct tests to detect mucormycosis [10]. Despite this, we have recently reported the development of species-specific [11], genus-specific [12] and pan-*Mucorales*-specific [13,14] monoclonal antibodies (mAbs) that detect *Mucorales* extracellular polysaccharides (EPSs), and their incorporation into lateral-flow immunoassays [11,13] for the detection of these antigenic biomarkers of mucormycosis in humans [15].

Here, we report the development of a murine IgG1 mAb, FH12, specific to EPSs produced during active growth of *Mucorales* pathogens. When incorporated into a Sandwich-Enzyme-Linked Immunosorbent Assay (Sandwich-ELISA), mAb FH12 is able to detect *Mucorales* EPSs in human serum with limits of detection of pg/mL. We show how the Sandwich-ELISA can be used to detect mucormycosis using tissue homogenates from a patient with a cutaneous and soft-tissue infection caused by *Lichtheimia ramosa*. This is the first time that a pan-*Mucorales*-specific Sandwich-ELISA has been reported for the detection of mucormycosis, enabling the laboratory-based detection of EPS antigens in human biofluids, which can act as signature molecules of invasive *Mucorales* infections.

## 2. Materials and Methods

### 2.1. Ethics Statement

Hybridoma work described in this study was conducted under a Home Office Project License, and was reviewed by the institution’s Animal Welfare Ethical Review Board (AWERB) for approval on 20 January 2022. The work was carried out in accordance with The Animals (Scientific Procedures) Act 1986 Directive 2010/63/EU, and followed all the Codes of Practice which reinforce this law, including all elements of housing, care, and euthanasia of the animals.

### 2.2. Fungal Culture

Fungi (Table 1 and Table 2) were routinely cultured on malt extract agar (MEA; 70145, Sigma, Poole, UK). The medium was autoclaved at 121 °C for 15 min prior to use, and fungi were grown at 30 °C. To induce sporulation of *Apophysomyces* and *Saksenaea* species, the methods described in Thornton and Davies [12] were used.

**Table 1.** Details of human pathogenic fungi used to determine the specificity of mAb FH12, and results of Direct-ELISA tests using purified EPS antigens. ^1^CBS; Westerdijk Fungal Biodiversity Institute, The Netherlands. CRT; C. R. Thornton, University of Exeter, UK. FGSC; Fungal Genetic Stock Centre, Kansas City University, USA. NCPF; National Centre for Pathogenic Fungi, UK. ^2^For DirectELISA tests, the absorbance values are the means of two replicate values ±SE, and the threshold absorbance value for test positivity is ≥0.100.

| Species | Isolate Number | Source<sup>1</sup> | Direct-ELISA (Abs 450nm)<sup>2</sup> |
| --- | --- | --- | --- |
| *Apophysomyces elegans* | 477.78 | CBS | 1.518±0.050 |
| *Apophysomyces mexicanus* | 136361 | CBS | 0.445±0.012 |
| *Apophysomyces ossiformis* | 125533 | CBS | 1.432±0.012 |
| *Apophysomyces variabilis* | 658.93 | CBS | 1.450±0.018 |
| *Aspergillus fumigatus* | Af293 | FGSC | 0.099±0.002 |
| *Cunninghamella bertholletiae* | 151.80 | CBS | 1.324±0.034 |
| *Fusarium solani* | 224.34 | CBS | 0.066±0.004 |
| *Lichtheimia corymbifera* | 109940 | CBS | 1.421±0.019 |
| *Lichtheimia hyalospora* | 146576 | CBS | 1.190±0.069 |
| *Lichtheimia ornata* | 142195 | CBS | 1.393±0.016 |
| *Lichtheimia ramosa* | 124197 | CBS | 1.507±0.007 |
| *Lichtheimia ramosa* | 112528 | CBS | 1.460±0.063 |
| *Lichtheimia ramosa* | 2845 | NCPF | 1.410±0.028 |
| *Lomentospora prolificans* | 3.1 | CRT | 0.074±0.006 |
| *Mucor circinelloides* | B5-2 | CRT | 1.545±0.031 |
| *Mucor racemosus* f. *racemosus* | 222.81 | CBS | 1.426±0.063 |
| *Rhizomucor pusillus* | 120587 | CBS | 1.327±0.016 |
| *Rhizopus arrhizus* | 111233 | CBS | 1.320±0.060 |
| *Rhizopus arrhizus* var. *arrhizus* | 112.07 | CBS | 1.346±0.022 |
| *Rhizopus microsporus* var. *rhizopodiformis* | 102277 | CBS | 1.550±0.044 |
| *Saksenaea vasiformis* | 113.96 | CBS | 0.566±0.018 |
| *Scedosporium aurantiacum* | 121926 | CBS | 0.088±0.001 |
| *Scedosporium boydii* | 835.96 | CBS | 0.084±0.001 |

**Table 2.** Details of fungi used in this study, and results of Sandwich-ELISA tests using culture filtrates from fungi grown as shake cultures for 72 h in YNB+G medium. ^1^CBS; Westerdijk Fungal Biodiversity Institute, The Netherlands. CRT; C. R. Thornton, University of Exeter, UK. FGSC; Fungal Genetic Stcok Centre, Kansas City University, USA. NCPF; National Centre for Pathogenic Fungi, UK. ^2^For Sandwich-ELISA tests, the absorbance values are the means of three replicate values ±SE, and the threshold absorbance value for test positivity is ≥0.100.

| Species | Isolate Number | Source<sup>1</sup> | Sandwich-ELISA (Abs 450nm)<sup>2</sup> |
| --- | --- | --- | --- |
| *Absidia glauca* | 2 | CRT | 0.802±0.007 |
| *Absidia spinosa* | 3 | CRT | 0.681±0.013 |
| *Actinomucor elegans* var. *kuwaitensis* | 117697 | CBS | 0.769±0.055 |
| *Apophysomyces elegans* | 477.78 | CBS | 0.625±0.034 |
| *Apophysomyces mexicanus* | 136361 | CBS | 0.891±0.004 |
| *Apophysomyces ossiformis* | 125533 | CBS | 0.827±0.052 |
| *Apophysomyces variabilis* | 658.93 | CBS | 0.790±0.038 |
| *Aspergillus fumigatus* | Af293 | FGSC | 0.087±0.014 |
| *Aspergillus flavus* | 144B | CRT | 0.065±0.020 |
| *Aspergillus nidulans* | A4 | FGSC | 0.075±0.009 |
| *Aspergillus niger* | 102.4 | CBS | 0.063±0.003 |
| *Aspergillus terreus* var. *terreus* | 601.65 | CBS | 0.057±0.009 |
| *Candida albicans* | SC5314 | SB | 0.048±0.002 |
| *Cryptococcus neoformans* | 8710 | CBS | 0.075±0.025 |
| *Cunninghamella bertholletiae* | 151.80 | CBS | 0.821±0.042 |
| *Fusarium oxysporum* | 167.3 | CBS | 0.035±0.002 |
| *Lichtheimia corymbifera* | 109940 | CBS | 0.835±0.020 |
| *Lichtheimia corymbifera* | 120580 | CBS | 0.853±0.020 |
| *Lichtheimia hyalospora* | 146576 | CBS | 0.867±0.009 |
| *Lichtheimia ornata* | 142195 | CBS | 0.845±0.026 |
| *Lichtheimia ramosa* | 124197 | CBS | 0.790±0.016 |
| *Lichtheimia ramosa* | 112528 | CBS | 0.790±0.016 |
| *Lichtheimia ramosa* | 2845 | NCPF | 0.777±0.074 |
| *Lomentospora prolificans* | 3.1 | CRT | 0.049±0.004 |
| *Mucor ardhlaengiktus* | 126271 | CBS | 0.682±0.005 |
| *Mucor circinelloides* | 124429 | CBS | 0.799±0.054 |
| *Mucor circinelloides* | B5-2 | CRT | 0.840±0.044 |
| *Mucor circinelloides* f. *circinelloides* | E2A | CRT | 0.776±0.046 |
| *Mucor circinelloides* f. *circinelloides* | 120582 | CBS | 0.757±0.035 |
| *Mucor indicus* | 120.08 | CBS | 0.734±0.005 |
| *Mucor irregularis* | 103.93 | CBS | 0.725±0.078 |
| *Mucor plumbeus* | 96 | CRT | 0.847±0.025 |
| *Mucor racemosus* f. *racemosus* | 112382 | CBS | 0.404±0.049 |
| *Mucor velutinosus* | 126272 | CBS | 0.725±0.036 |
| *Phycomyces nitens* | 133 | CRT | 0.709±0.013 |
| *Rhizomucor pusillus* | 120586 | CBS | 0.731±0.022 |
| *Rhizomucor pusillus* | 120587 | CBS | 0.500±0.026 |
| *Rhizopus arrhizus* | TV4 | CRT | 0.751±0.089 |
| *Rhizopus arrhizus* | 2634 | CBS | 0.570±0.095 |
| *Rhizopus arrhizus* var. *arrhizus* | 112.07 | CBS | 0.405±0.004 |
| *Rhizopus arrhizus* var. *arrhizus* | 118614 | CBS | 0.644±0.032 |
| *Rhizopus arrhizus* var. *delemar* | 2601 | NCPF | 0.606±0.014 |
| *Rhizopus arrhizus* var. *delemar* | 607.68 | CBS | 0.313±0.004 |
| *Rhizopus arrhizus* var. *delemar* | 544.80 | CBS | 0.677±0.039 |
| *Rhizopus azygosporus* | 357.93 | CBS | 0.533±0.001 |
| *Rhizopus homothallicus* | 336.62 | CBS | 0.481±0.004 |
| *Rhizopus microsporus* var. *rhizopodiformis* | 102277 | CBS | 0.650±0.088 |
| *Rhizopus microsporus* var. *rhizopodiformis* | 220.92 | CBS | 0.727±0.016 |
| *Rhizopus microsporus* var. *rhizopodiformis* | 118987 | CBS | 0.612±0.073 |
| *Rhizopus schipperae* | 138.95 | CBS | 0.474±0.014 |
| *Rhizopus stolonifer* var. *stolonifer* | 389.95 | CBS | 0.734±0.040 |
| *Saksenaea erythrospora* | 138279 | CBS | 0.792±0.092 |
| *Saksenaea vasiformis* | 113.96 | CBS | 0.703±0.050 |
| *Scedosporium apiospermum* | 117407 | CBS | 0.056±0.011 |
| *Scedosporium aurantiacum* | 118934 | CBS | 0.070±0.005 |
| *Scedosporium aurantiacum* | 121926 | CBS | 0.071±0.006 |
| *Scedosporium boydii* | 835.96 | CBS | 0.038±0.002 |
| *Scedosporium boydii* | 100870 | CBS | 0.075±0.012 |
| *Scedosporium boydii* | 100393 | CBS | 0.083±0.007 |
| *Scedosporium dehoogii* | 117406 | CBS | 0.067±0.003 |
| *Scedosporium fusoideum* | 106.53 | CBS | 0.064±0.003 |
| *Syncephalastrum racemosum* | 155 | CRT | 0.765±0.035 |
| YNB+G only | - | - | 0.067±0.001 |

### 2.3. Production of Hybridomas and Screening by ELISA

#### 2.3.1. Hybridoma Production

Extracellular polysaccharides (EPSs) were prepared as described elsewhere [11—13]. For hybridoma production, the immunogen comprised a 1 mg/mL solution of EPS from *Apophysomyces variabilis* CBS658.93, with six-week-old BALB/c white mice each given four intra-peritoneal injections (300 µL per injection) of immunogen at 2-wk intervals, and a single booster injection 5 d before fusion. Hybridoma cells were produced by the method described elsewhere, and monoclonal antibody (mAb)-producing clones identified in Indirect-ELISA tests.

#### 2.3.2. Indirect-Enzyme-Linked Immunosorbent Assay

For Indirect-ELISA, EPS at a concentration of 20 μg/mL phosphate-buffered saline (PBS; 137 mM NaCl, 2.7 mM KCl, 8 mM Na_2_HPO_4_, 1.5 mM KH_2_PO_4_ [pH 7.2]) immobilized to the wells of Maxisorp microtiter plates (Nunc) at 50 µL/well. Wells containing immobilized antigen were incubated with 50 µL of mAb hybridoma tissue culture supernatant (TCS) for 1 h, after which wells were washed three times, for 5 min each, with PBST (PBS containing 0.05% (vol:vol) Tween-20). Goat anti-mouse polyvalent immunoglobulin (G, A, M) peroxidase conjugate (PA1-84388; Invitrogen, Loughborough, UK), diluted 1:5000 (vol:vol) in PBST, was added to the wells and incubated for a further hour. The plates were washed with PBST as described, given a final 5 min wash with PBS, and bound antibody visualised by incubating wells with tetramethyl benzidine (TMB) substrate solution for 30 min, after which reactions were stopped by the addition of 3 M H_2_SO_4_. Absorbance values were determined at 450 nm using a microplate reader (infinite F50, Tecan Austria GmbH, Reading, UK). Control wells were incubated with tissue culture medium (TCM) containing 10% (vol:vol) foetal bovine serum (F7254, Sigma, Poole, UK) only. All incubation steps were performed at 23 °C in sealed plastic bags. The threshold for detection of the antigen in Indirect-ELISA was determined from control means (2 x TCM absorbance values). These values were consistently in the range of 0.050-0.100. Consequently, absorbance values ≥0.100 were considered as positive for the detection of antigen.

### 2.4. Determination of Ig Class and Sub-cloning Procedure

The Ig class of mAbs was determined by using antigen-mediated ELISA. Wells of three replicate microtiter plates coated with 20 μg EPS/mL PBS were incubated successively with hybridoma TCS for 1 h; goat anti-mouse IgG1-, IgG2a-, IgG2b-, IgG3-, IgM- or IgA-specific antiserum (ISO-2, Sigma) diluted 1:3000 (vol:vol) in PBST for 30 min, and rabbit anti-goat peroxidase conjugate (A5420, Sigma, Poole, UK) diluted 1:1000 (vol:vol) for a further 30 min. Bound antibody was visualised with TMB substrate as described. Hybridoma cell lines were sub-cloned three times by limiting dilution, and cell lines were grown in bulk in a non-selective medium, preserved by slowly freezing in FBS/dimethyl sulfoxide (92:8 vol:vol), and stored in liquid N_2_.

### 2.5. Antibody Purification and Enzyme Conjugation

Hybridoma TCS of mAb FH12 was harvested by centrifugation at 2,150 x *g* for 40 min at 4 °C, followed by filtration through a 0.8 µM cellulose acetate filter (10462240, GE Healthcare Life Sciences, Little Chalfont, UK). Culture supernatant was loaded onto a HiTrap Protein A column (17-0402-01, GE Healthcare Life Sciences, Little Chalfont, UK) using a peristaltic pump P-1 (18-1110-91, GE Healthcare Life Sciences, Little Chalfont, UK) with a low pulsation flow of 1 mL/min. Columns were equilibrated with 10 mL of PBS and column-bound antibody was eluted with 5 mL of 0.1 M glycine-HCl buffer (pH 2.5) with a flow rate of 0.5 mL/min. The buffer of the purified antibody was exchanged to PBS using a disposable PD-10 desalting column (17-0851-01, GE Healthcare Life Sciences, Little Chalfont, UK). Following purification, the antibody was sterile-filtered with a 0.24 µm syringe filter (85037-574-44, Sartorius, Göttinghen, Germany) and stored at 4 °C. Protein concentrations were determined using a Nanodrop spectrophotometer with protein concentration calculated using the mass extinction coefficient of 13.7 at 280 nm for a 1% (10 mg/mL) IgG solution. Antibody purity was confirmed by SDS-PAGE and gel staining using Coomassie Brilliant Blue R-250 dye (Thermo Fisher Scientific, Loughborough, UK). Protein A-purified mAb FH12 was conjugated to horseradish peroxidase (HRP) for ELISA studies using a Lightning-Link horseradish peroxidase conjugation kit (701-0000, Bio-Techne Ltd., Abingdon, UK), or to alkaline phosphatase (AKP) for western blotting studies using a Lightning-Link alkaline phosphatase conjugation kit (702-0010, Bio-Techne Ltd., Abingdon, UK).

### 2.6. Determination of Antibody Specificity

For antibody specificity tests, wells of two replicate plates were coated with purified EPS antigens at a concentration 20 μg/mL PBS from different *Mucorales* fungi and from unrelated moulds of clinical importance (Table 1). The wells were assayed by Direct-ELISA using the FH12-HRP conjugate diluted 1:5000 (vol:vol) in PBST for 1 h, followed by TMB substrate solution for 30 min.

### 2.7. Epitope Characterisation by Periodate Oxidation and Heat Treatment

For periodate oxidation, three replicate microtiter plates containing immobilised EPS from *Apophysomyces variabilis* strain CBS658.93 at a concentration of 20 μg/mL PBS were incubated with 50 μL of sodium *meta*-periodate solution (20 mM NaIO_4_ in 50 mM sodium acetate buffer (pH4.5)) or acetate buffer only (control) for 24, 4, 3, 2, 1, or 0 h, at 4 °C in sealed plastic bags. The plates were given four 3 min washes with PBS before assay by Direct-ELISA using the FH12-HRP conjugate as described. The heat stability of the FH12 epitope was determined by heating three replicate EPS samples from *Apophysomyces variabilis* strain CBS658.93 at a concentration of 20 μg/mL PBS in a boiling water bath. At 5 min, and 10 min intervals, 50 μL volumes were removed and, after cooling, were transferred to the wells of microtiter plates for assay by Direct-ELISA using the FH12-HRP conjugate as described.

### 2.8. Polyacrylamide Gel Electrophoresis and Western Blotting

Sodium-dodecyl-sulphate-polyacrylamide gel electrophoresis (SDS-PAGE) was carried out using 4–20% gradient polyacrylamide gels (161-1159, Bio-Rad, Watford, UK) under denaturing conditions. Antigens were separated electrophoretically at 165 V and prestained markers (161-0318, Bio-Rad, Watford, UK) were used for molecular weight determinations. For western blotting, separated antigens were transferred electrophoretically onto a PVDF membrane (162-0175, Bio-Rad, Watford, UK) for 2 h at 75 V, and the membrane was blocked for 16 h at 4 °C in PBS containing 1% (wt:vol) bovine serum albumin (BSA). Blocked membranes were incubated with FH12-AKP conjugate diluted 1:5,000 (vol:vol) in PBS containing 0.5% (wt/vol) BSA for 2 h at 23 °C. Membranes were washed three times with PBS, once with PBST and bound antibody visualised by incubation in substrate solution. Reactions were stopped by immersing membranes in dH_2_O, and membranes were then air dried between sheets of Whatman filter paper.

### 2.9. Spatio-Temporal Production of EPS Antigens In Vitro

#### 2.9.1. Colony Immunoblotting

For colony immunoblots [13], MEA was inoculated centrally with 5 mL of a 10^3^ spores/mL spore suspension of *Apophysomyces variabilis* strain CBS658.93 and incubated for 16 h at 30 °C, after which colonies were overlayed with PVDF membrane (162-0175, Bio-Rad, Watford, UK) for 8 h to bind extracellular antigens. The membranes were removed and discarded, the colonies incubated for a further 16 h, and the blotting procedure repeated. The membranes were blocked and processed with FH12-AKP conjugate as described for western blotting, with mAb TG11-AKP conjugate [12] diluted 1:5,000 (vol:vol) used as the control.

#### 2.9.2. Immunofluorescence

Immunofluorescence was carried out as described elsewhere [16] using spores of *Mucor circinelloides* f. *circinelloides* CBS120582 germinated on glass slides at 30 °C in autoclaved YNB+G liquid medium [10*—*12], and with a 1:500 (vol:vol) dilution in PBS of mAb FH12 directly conjugated to the fluorophore Cy3 using a Lightning-Link (R) Rapid Cy3 conjugation kit (340-0030, Bio-Techne Ltd., Abingdon, UK). Epifluorescence imaging was conducted with a SP8 confocal laser microscope under 550 nm absorbance and 570 nm emission spectra.

#### 2.9.3. Shake Culture

Conical flasks (250 mL) each containing 100 mL of autoclaved YNB+G liquid medium [11—13] were inoculated with spores of *Mucorales* fungi and unrelated moulds of clinical importance (Table 2) to a final concentration of 10^3^ spores/mL. The cultures were incubated at 30 °C with shaking (120 RPM) in a New Brunswick orbital shaker. At 24 h intervals, three replicate flasks were harvested, the culture fluids separated from mycelium by filtration through Miracloth (475855-1R, Merck Millipore, Germany), and then stored at -20 °C. Mycelial biomass was dried for 4 d at 80 °C and weighed. On thawing, culture fluids were centrifugation for 5 min at 16,000 x g, and then assayed by Sandwich-ELISA and by western blotting.

### 2.10. Sandwich-ELISA

The Sandwich-ELISA procedure is illustrated in Figure 1B. Wells of Maxisorp microtiter plates (Nunc) were coated with 50 µL volumes of Protein A-purified mAb FH12 at a concentration of 4 μg/mL PBS. After incubation for 16 h at 4 °C, the wells were washed three times (5 min each wash) with PBST, once with PBS for 5 min, and then given a final rinse with dH_2_O before air-drying at 23 °C. Antibody-coated wells were blocked for 15 min with 100 μL of PBS containing 1.0% (wt:vol) BSA, and then incubated at 23 °C for 1 h with 50 µL of culture filtrates diluted 1:1 (vol:vol) with PBST (control wells incubated with YNB+G medium only diluted 1:1 (vol:vol) with PBST). The wells were given four 5-min washes with PBST, and then incubated for 50 min at 23 °C with FH12-HRP conjugate diluted 1 in 10,000 (vol:vol) in PBST containing 0.25% (wt:vol) BSA. The wells were washed four times with PBST as described, given a final 5-min wash with PBS, and bound antibody visualised by incubating wells with tetramethyl benzidine (TMB) substrate solution for 15 min. Enzyme-substrate reactions were stopped by the addition of 3 M H_2_SO_4_, and absorbance values were determined at 450 nm using a microplate reader (infinite F50, Tecan Austria GmbH, Reading, UK). All incubation steps were performed at 23 °C in sealed plastic bags.

### 2.11. Detection of EPS Antigens in Human Serum and Limits of Detection of Sandwich-ELISA

Normal serum from healthy AB blood group males (H6914, Sigma, Poole, UK) was spiked with purified EPS from *Apophysomyes variabilis* CBS658.93, *Cunninghamella bertholletiae* CBS151.80, *Mucor circinelloides* B5-2, *Lichtheimia corymbifera* CBS109940, *Lichtheimia ramosa* CBS112528, *Rhizomucor pusillus* CBS120587, *Rhizopus arrhizus* var. *arrhizus* CBS112.07, *Rhizopus microsporus* var. *rhizopodiformis* CBS102277, and stored as aliquots at -20 °C prior to use. The standard operating procedure for serum pre-treatment and testing using the Sandwich-ELISA is illustrated in Figure 1C. On thawing, 50 μL spiked or control (unspiked) serum was mixed 1:2 (vol:vol) with PBS containing 0.5% (wt:vol) Na_2_-EDTA (D/0700/53, Thermo Fisher Scientific, Loughborough, UK)(pH6.0), and heated in a boiling water bath for 3 min. The heated mixture was centrifuged at 16,000 x *g* for 5 min, the clear supernatant mixed 1:1 (vol:vol) with PBST, and the resultant solution assayed by Sandwich-ELISA (50 μL per well) as described.

#### 2.12. Detection of Cutaneous Mucormycosis

Tissue homogenates from an immunocompetent male patient with cutaneous and soft tissue mucormycosis of the right hand caused by the *Mucorales* pathogen *Lichtheimia ramosa* strain 266533346 [17], were used to evaluate the Sandwich-ELISA as a diagnostic immunoassay to detect human mucormycosis. The tissue homogenates in dH_2_0 were centrifuged at 17,000 x *g* for 5 min to remove cellular debris, and undiluted 50 μL samples of supernatants tested in the Sandwich-ELISA (Figure 1B) as described. Sterile dH_2_0 acted as the control.

### 2.13. Statistical Analysis

Numerical data were analysed using the statistical programme Minitab (Minitab 16; Minitab, Coventry, UK). Analysis of variance (ANOVA) was used to compare means, and post hoc Tukey–Kramer analysis was then performed to determine statistical significance.

## 3. Results

### 3.1. Production of Hybridomas and mAb Isotyping

A single hybridoma fusion was performed, and 327 hybridoma cell lines were tested in indirect ELISA tests for recognition of the immunogen. Seven cell lines produced EPS-reactive antibodies, 5 of which produced mAbs of the immunoglobulin class, G1 (IgG1). The remaining 2 cell lines produced mAbs of the class, G2b (IgG2b). The cell line FH12 (an IgG1) was selected for further evaluation due to its specificity for *Apophysomyces* species, and lack of cross-reactivity with non-*Mucorales* fungi.

### 3.2. Specificity of mAb FH12 and Epitope Characterisation

In Direct-ELISA tests of purified EPSs from the most common *Mucorales* and non-*Mucorales* mould pathogens of humans, mAb FH12 was found to be pan-*Mucorales*-specific, reacting with all of the *Mucorales* fungi tested (Table 1). No cross-reactivity was found with the unrelated mould pathogens of clinical importance (*Aspergillus, Fusarium, Lomentospora*, and *Scedosporium* species) [18]. Direct-ELISA tests showed that mAb FH12 binds to a periodate-insensitive epitope (Figure 2A), with no loss in mAb binding due to periodate treatment over the 24 h experimental period. Heating of the EPS antigen led to a progressive loss in antibody binding compared to the control (no heat treatment; 0 min) over the 60 min experimental period. However, the loss in mAb binding, determined by successive decreases in ELISA absorbance values compared to the control, was significant (*p* <0 .05) at time points beyond the first 10 min of heating only (Figure 2B).

**Figure 2.**
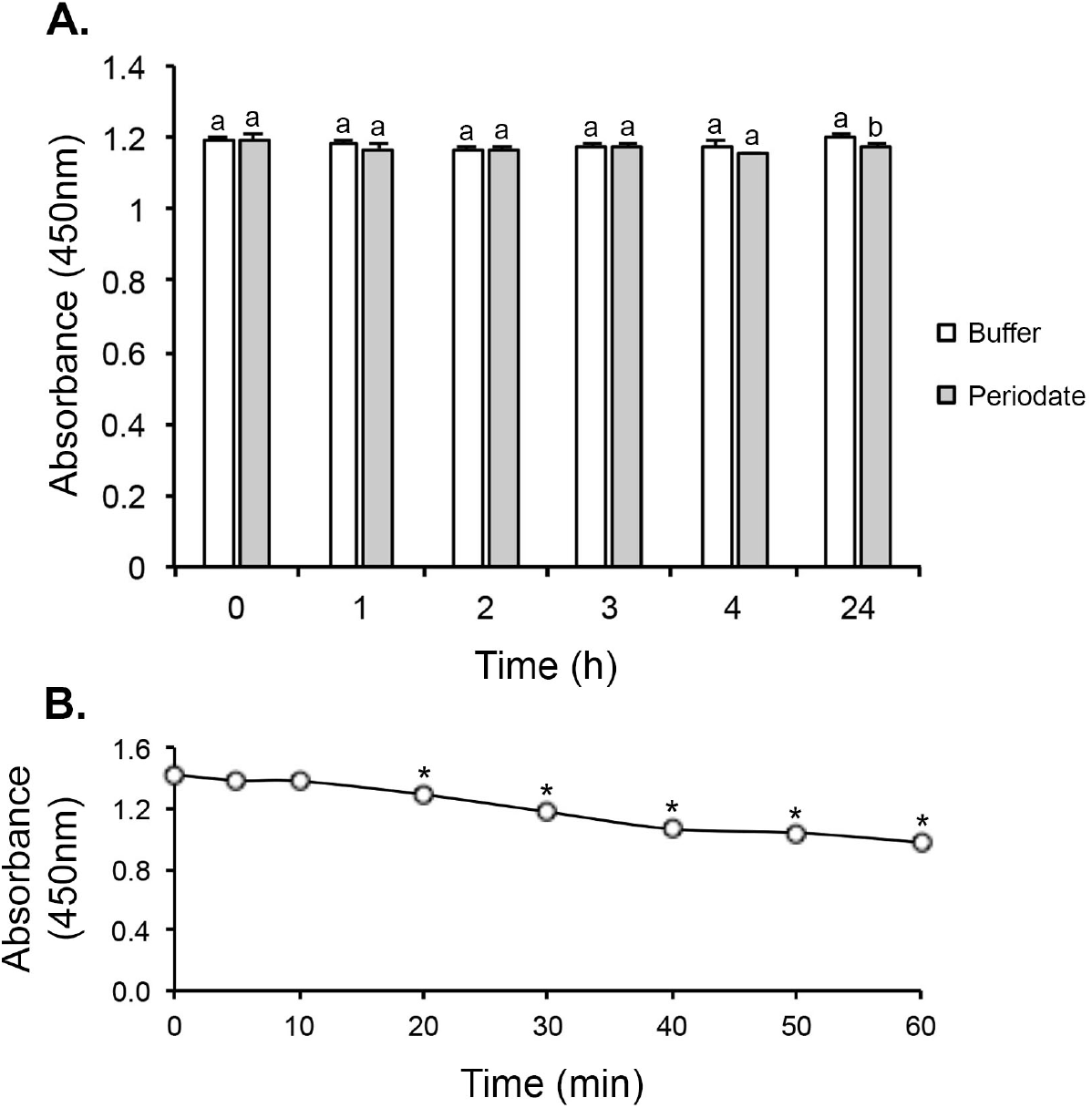
Epitope characterisation by periodate oxidation and heat treatment. (**A**) Effect of periodate oxidation on mAb FH12 binding in Direct-ELISA to EPS from *Apophysomyces variabilis* CBS658.93. There was no significant effect of periodate treatment (shaded bars) compared to the control (open bars) over the 24 h period of treatment. (**B**) Effect of heat treatment at 100 °C on binding of mAb FH12 to EPS from *Apophysomyces variabilis* CBS658.93. There was no significant effect on mAb binding for 5 min or 10 min heating compared to the control (no heat treatment; 0 min), but there were successive and significant decreases in mAb binding between 20 min and 60 min compared to the control. For both the periodate and heat treatments, bars and data points are the means of 3 replicates ± SE, and bars (**A**) with the same letters show no significant differences (*p* < 0.05) between periodate treatment and matched controls (buffer only) at each time point. Data points (**B**) with an asterisk * show significant differences (Student’s *t*-test (*p* < 0.05)) between heat treatment and the control (no heat treatment; 0 min) at each time point.

### 3.3. Colony Immunoblotting and Immunofluorescence

In colony immunoblots, production of the FH12 antigen was shown to be extracellular and was associated with the growing margins of the fungal colony (Figure 3A). This staining pattern was similar to the staining pattern found with the pan-*Mucorales*-specific IgG1 control mAb TG11 (Figure 3B). In immunofluorescence studies of germinated cells of *Mucor circinelloides* f. *circinelloides* CBS120582, the FH12 antigen was localised to the outer cell wall of hyphae and germinated spores (Figure 3C and Figure 3E), but was not found on the outer cell wall of the ungerminated conidium (indicated by the asterisk * in Figure 3F).

**Figure 3.**
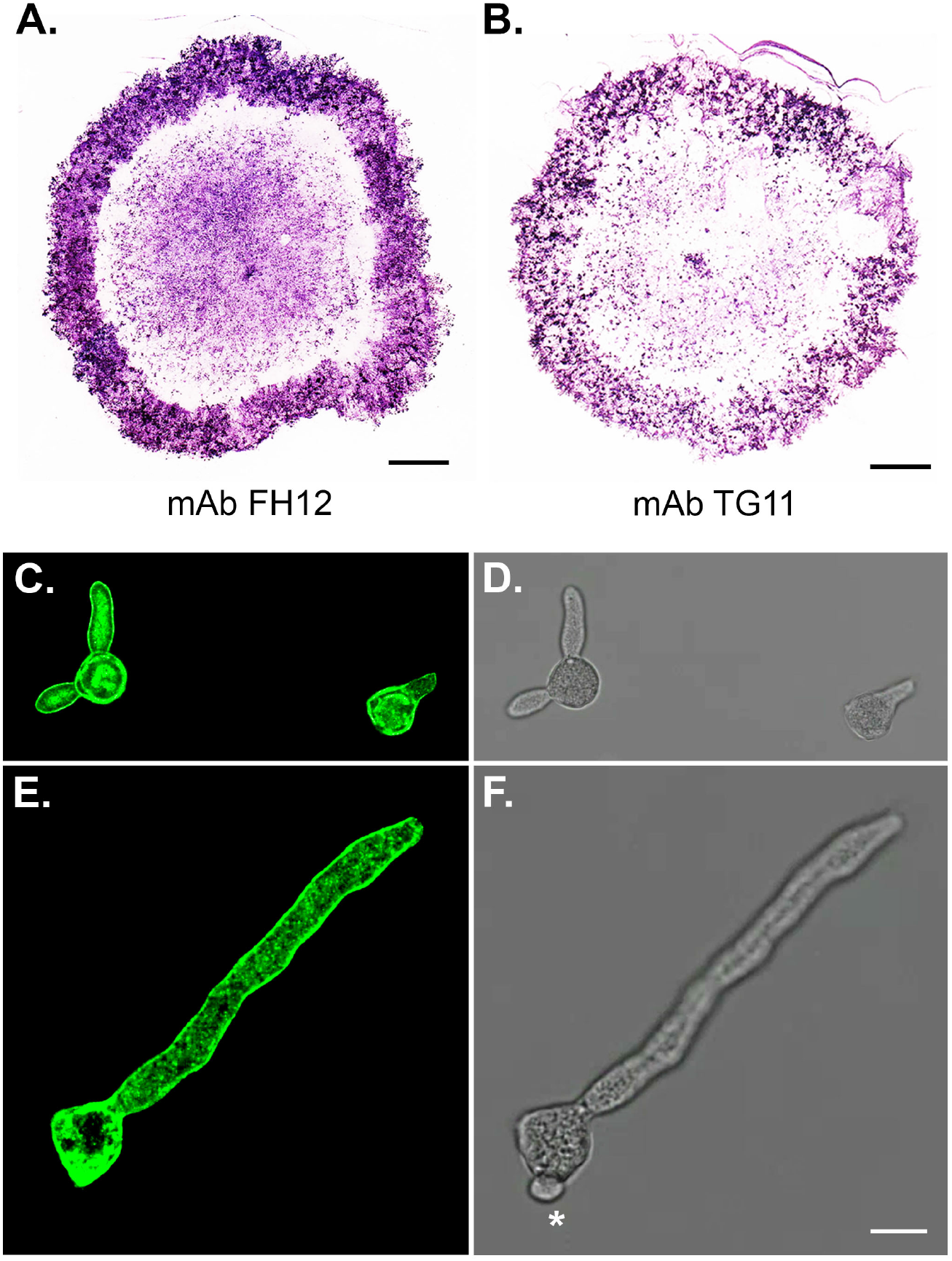
Colony immunoblots and immunofluorescence imaging of the *Mucorales* pathogens *Apophysomyces variabilis* CBS658.93, and *Mucor circinelloides* f. *circinelloides* CBS120582. (**A**) Colony immunoblots of *Apophysomyces variabilis* CBS658.93, showing extracellular production of mAb FH12 and mAb TG11 (Thornton et al., 2023) EPS antigens. Note the intense immuno-staining of extracellular antigens produced at the colony margins during active growth of the pathogen. Scale bars = 0.5 cm. (**C**,**E**) Epifluorescence images of germinated spores of *Mucor circinelloides* f. *circinelloides* CBS120582 immuno-stained with mAb FH12-Cy3 conjugate, showing intense immunofluorescence of the hyphal and spore cell walls of germinated conidiospores. The corresponding brightfield images are shown in (**D**) and (**F**), respectively. Note lack of immunofluorescence of ungerminated spore in (**E**), indicated by asterisk * in corresponding brightfield image (**F**). Scale bar = 7 μm.

**Figure 3.**
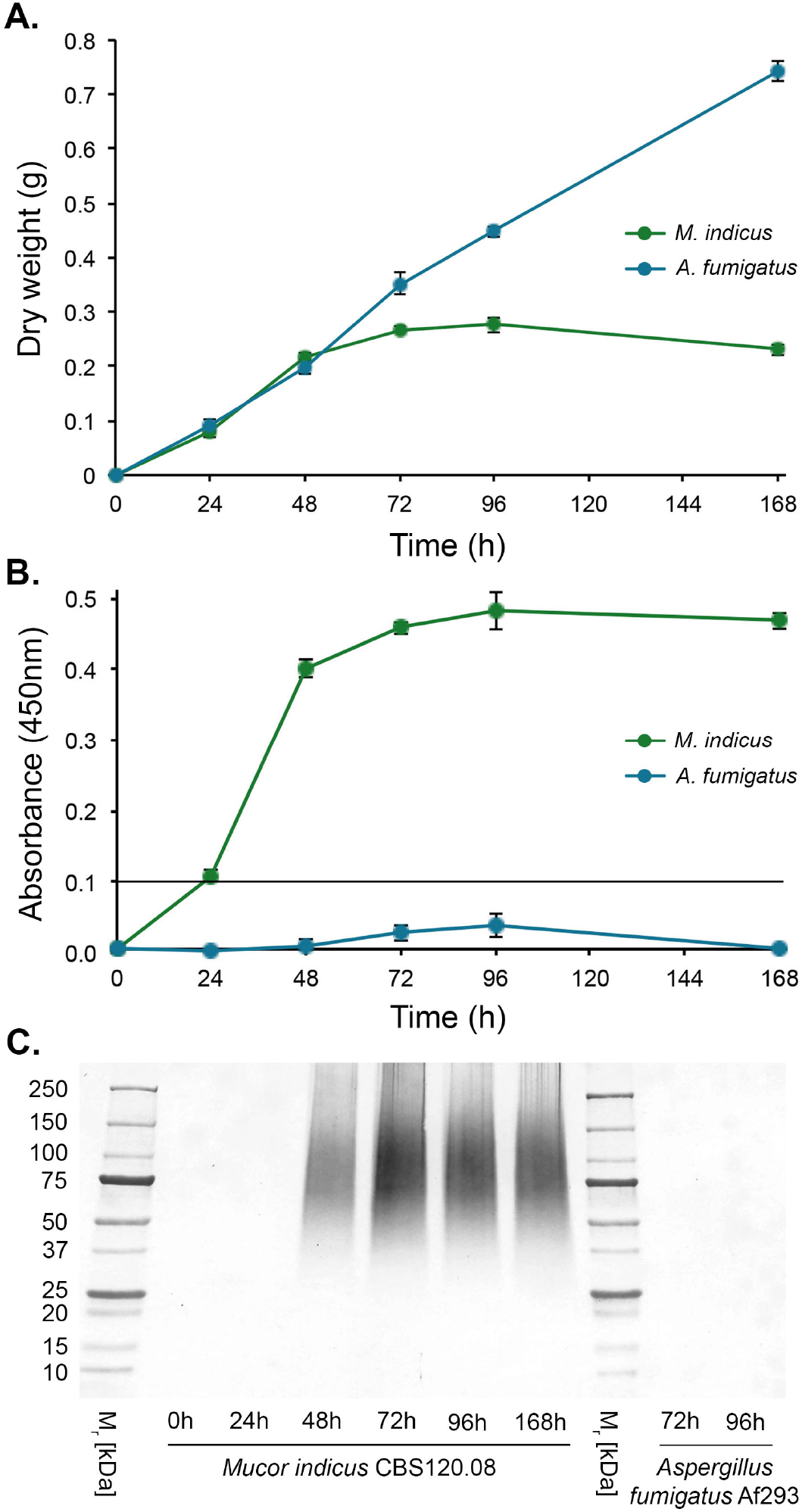
Production of the FH12 EPS antigen by the *Mucorales* pathogen, *Mucor indicus* CBS120.08. **(A)**. Dry weights of the pathogen over the 168 h experimental period. **(B)** Sandwich-ELISA of YNB+G culture filtrates. Each data point is the mean of three replicates ± SE **(B**,**C)**. The threshold absorbance value for detection of the antigen in the Sandwich-ELISA is 0.1 (indicated by the line in **B**), with values above this threshold positive for the detection of the FH12 antigen. **(C)** Western blot of culture filtrates using mAb FH12, showing the presence of immunoreactive EPS antigens with molecular weights of between ∼25 kDa and ∼250 kDa which are absent in the 72 h and 96 h culture filtrates of the unrelated human pathogen, *Aspergillus fumigatus* Af293.

### 3.4. Sandwich-ELISA and Western Blotting of Cultures Filtrates

Using culture filtrates from yeasts and moulds grown for 72 h in YNB+G liquid medium, the Sandwich-ELISA was shown to be pan-*Mucorales*-specific (Table 2), reacting strongly with filtrates from *Mucorales* fungi only (Sandwich-ELISA absorbance values >0.100, the threshold absorbance value for test positivity). There was no cross-reaction of mAb FH12 with filtrates from unrelated yeasts and moulds of clinical importance including *Aspergillus* spp., *Candida albicans, Cryptococcus neoformans, Fusarium* spp., *Lomentospora prolificans*, and *Scedosporium* spp. (all absorbance values <0.100, the threshold for test positivity).

A study of antigen production in YNB+G shake culture by *Rhizomucor pusillus* CBS120587, a *Mucorales* pathogen responsible for gastro-intestinal mucormycosis in humans, showed that growth of the pathogen (dry weight (g) of hyphal biomass) plateaued between 72 h and 96 h, while growth of the unrelated mould pathogen *Aspergillus fumigatus* Af2923 increased up to the final sampling point at 168 h (Figure 3A). Sandwich-ELISA tests of the culture filtrates showed that the FH12 EPS antigen was secreted into the culture medium and was first detectable in the Sandwich-ELISA 24 h post-inoculation (Figure 3B). Antigen production continued to rise and, similar to hyphal biomass, plateaued between 72 h and 96 h post-inoculation. Western blots of culture filtrates showed that mAb FH12 reacted strongly with immunoreactive EPS antigens with molecular weights of between ∼25 kDa and ∼250 kDa (Figure 3C), which were first detectable at 48 h post-inoculation. The antibody did not cross-react with 72 h old and 96 h old culture filtrates of *Aspergillus fumigatus* Af293 (Figure 3C).

### 3.5. Sandwich-ELISA of Human Serum and Limits of Detection

Using running buffer (RB) spiked with purified *Mucorales* EPSs (Table 3), the Sandwich-ELISA was shown to have limits of detection of pg EPS/mL (*Apophysomyces variabilis, Cunninghamella bertholletiae, Lichtheimia corymbifera, Mucor circinelloides*, and *Rhizomucor pusillus*) and in the ng EPS/mL range (*Rhizopus arrhizus* and *Rhizopus microsporus*). The Sandwich-ELISA is compatible with human serum (HS), with limits of detection in the pg/mL serum range (*A. variabilis, C. bertholletiae, M. circinelloides*, and *R. pusillus*) and in the ng/mL range (*Lichtheimia corymbifera, Rhizopus arrhizus*, and *Rhizopus microsporus*).

**Table 3.** Limits of Detection (LOD) of the Sandwich-ELISA in ng/mL using running buffer (RB) or human serum (HS) spiked with purified EPSs from *Mucorales* fungi. The LOD values are determined from the mean absorbance values of 4 replicate samples showing significant differences (Student’s t-test (*p* < 0.05)) compared to matched replicate controls (RB only or normal serum only) in each dilution series.

| *Mucorales* Species | Isolate Number | Limit of Detection in RB (ng/mL) | Limit of Detection in HS (ng/mL) |
| --- | --- | --- | --- |
| *Apophysomyces variabilis* | 658.93 | 0.45 | 0.57 |
| *Cunninghamella bertholletiae* | 151.80 | 0.07 | 0.35 |
| *Lichtheimia corymbifera* | 109940 | 0.46 | 4.59 |
| *Lichtheimia ramosa* | 124197 | 3.23 | 1.01 |
| *Mucor circinelloides* | B5-2 | 0.44 | 0.88 |
| *Rhizomucor pusillus* | 120587 | 0.19 | 0.24 |
| *Rhizopus arrhizus* var. *arrhizus* | 112.07 | 36.13 | 72.25 |
| *Rhizopus microsporus* var. *rhizopodiformis* | 102277 | 10.20 | 65.25 |

### 3.6. Detection of Cutaneous Mucormycosis

In Sandwich-ELISA tests, the soluble FH12 EPS antigen was readily detectable in tissue homogenates from a cutaneous and soft tissue mucormycosis caused by the *Mucorales* pathogen *Lichtheimia ramosa* strain 266533346 (Figure 5) recovered during surgical debridement of the patient’s hand on day 5 (D5) and day 14 (D14) of hospital admission. The mean absorbance values for the D5 (1.806±0.076) and D14 (0.157±0.006) tissue homogenate samples were significantly greater than the mean value for the control (0.065±0.004), showing the presence of the *Mucorales*-specific FH12 antigen in the homogenate samples.

**Figure 5.**
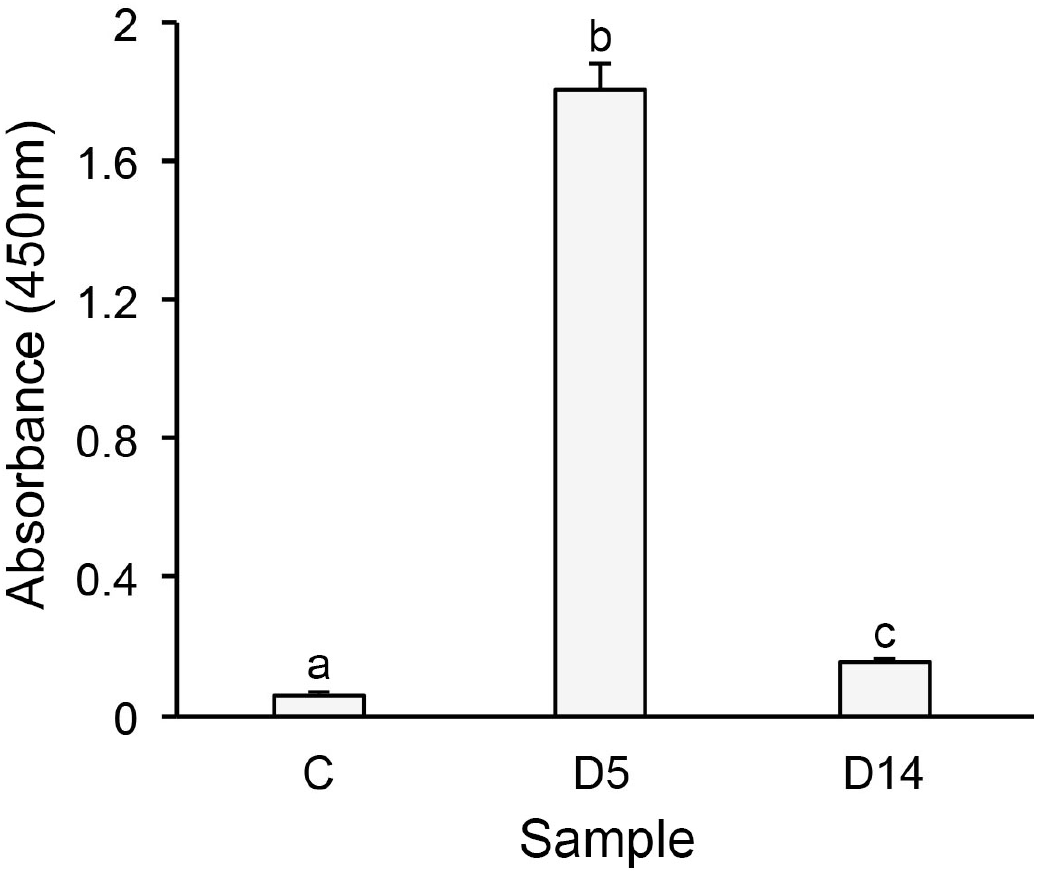
Detection of cutaneous mucormycosis by Sandwich-ELISA. The mean absorbance values for the day 5 (D5) and day 14 (D14) tissue homogenate samples are significantly greater than the mean absorbance value for the negative control comprising sterile dH^2^O (C), showing the presence of soluble *Mucorales*-specific FH12 antigen in the homogenate samples. Bars are the means of 3 technical replicates ± SE, and bars with different letters show significant differences in absorbance values between samples (*p* < 0.05).

## 4. Discussion

In this paper, we describe the development of a pan-*Mucorales*-specific IgG1 monoclonal antibody (mAb), FH12, and a Sandwich-ELISA for the detection of *Mucorales*-specific extracellular polysaccharides (EPSs). While an indirect ELISA has been reported for the serological detection of mucormycosis using preparations of heterogeneous antigens from *Rhizopus arrhizus* [19], this is the first time, to our knowledge, that a mAb-based Sandwich-ELISA has been reported for the diagnosis of mucormycosis in humans.

As shown by immunoblotting of growing colonies and immunofluorescence of germinating spores, the EPSs bound by mAb FH12 are excreted by *Mucorales* fungi during active hyphal growth, while western blotting studies of cultures filtrates demonstrate production of EPSs with molecular weights (∼25 kDa to ∼250 kDa) similar to the *Mucorales* cell wall EPSs bound by mAb TG11 [13,14]. However, unlike mAb TG11 which binds to a single epitope present in *Mucorales* EPSs, thereby restricting its use to a Competitive format lateral-flow immunoassay [13], the repeat epitopes bound by mAb FH12 enable its use in a Sandwich format immunoassay (a Sandwich-ELISA). We have shown that the epitope bound by mAb FH12 is insensitive to mild periodate oxidation at acid pH, a modifying procedure commonly used for detecting anti-carbohydrate mAbs when used in conjunction with ELISA [20]. However, not all carbohydrate epitopes bound by mAbs are sensitive to periodate cleavage, particularly those dependent on a core trisaccharide rather than terminal residues. Consequently, we conclude that mAb FH12 binds to a core carbohydrate epitope rather than terminal sugar moieties in *Mucorales* EPSs.

Of the different types of ELISA format, the Sandwich format is the most sensitive [21], and forms the basis of commercial ELISA tests for the detection of invasive pulmonary aspergillosis using human serum and bronchoalveolar lavage fluid (BALf) samples [22,23]. The pan-*Mucorales*-specific Sandwich-ELISA described here is similarly compatible with human serum. The moderate heat stability of the FH12 epitope enables the heat treatment of serum with EDTA to prevent immunoassay prozone effects, enabling detection of *Mucorales* EPS antigens in spiked human serum with detection in the pg/mL to low ng/mL range. This range of sensitivity is similar to that of enzyme immunoassays for aspergillosis detection [22,23].

The specificity of mAb FH12 enables the differentiation of pathogenic *Mucorales* species from other moulds and yeasts (*Candida albicans* and *Cryptococcus neoformans*) of clinical importance. This is especially significant in mixed species infections with *Aspergillus* species [24], the cause of invasive pulmonary aspergillosis and most common differential diagnosis with mucormycosis, and where other moulds such as *Fusarium, Lomentospora*, and *Scedosporium* are present in polymicrobial infections [25—30].

While the FH12 Sandwich-ELISA has yet to be evaluated with clinical serum and BALf specimens, we were able to demonstrate its clinical utility to detect invasive *Mucorales* infection in a patient with skin and soft-tissue mucormycosis caused by *Lichtheimia ramosa* following a farming injury [17]. The soluble EPS biomarker bound by mAb FH12 was readily detectable in fluids from homogenised tissue removed during an early-stage debridement (day 5) of the infected wound. The biomarker was still detectable on day 14 of the infection, albeit significantly reduced compared to day 5, which was likely due to treatment with the anti-fungal drug amphotericin B and cessation of active *Mucorales* infection [17]. The results with the FH12 Sandwich-ELISA mirror those obtained with the TG11-LFD test [17], confirming the utility of *Mucorales* EPSs as signature molecules of mucormycosis in humans. Importantly, the Sandwich-ELISA was able to detect the EPS biomarker days before conventional microbiological tests (culture and histopathology of tissue biopsies) confirmed cutaneous infection by the *Mucorales* pathogen [17].

In conclusion, we have developed a sensitive and specific Sandwich-ELISA for the detection of mucormycosis, which has the potential to be used as a laboratory-based adjunct test for *Mucorales* infections in humans. A commercial EIA kit for the diagnosis of mucormycosis is currently in development.

## 5. Trademark

The word, *Zygo*Dx^®^ (EU018696066), is protected by ISCA Diagnostics Ltd. through the European Union Intellectual Property Office (EUIPO).

## Data Availability

All data produced in the present work are contained in the manuscript.

## Author Contributions

Conceptualisation, CRT and GED; methodology, CRT and GED; investigation, CRT and GED; formal analysis, CRT and GED; resources, CRT; data curation, CRT and GED; writing - original draft preparation, CRT; writing, review and editing, CRT and GED; supervision, CRT; project administration, CRT; funding acquisition, CRT. The authors have read and agreed to the content of the manuscript.

## Funding

This work was funded by ISCA Diagnostics Ltd. (project title: A Monoclonal Antibody and Sandwich-ELISA for Mucormycosis Detection).

## Acknowledgements

The authors are grateful to Dr Alyssa C. Hudson for the supply of remnant clinical samples, for the purposes of diagnostic testing under patient consent.

## Informed Consent Statement

Human tissue samples were obtained for diagnostic purposes with informed consent. Samples were tested, examined and disposed of, with informed written consent and in accordance with the Human Tissue Act 2004, and the Human Tissue Authority Code of Practice.

## Data Availability Statement

The data presented in this study are available on request from the corresponding author, but are not publicly available due to commercial confidentialities.

## Conflicts of Interest

CRT is Founder Director and an employee of ISCA Diagnostics Ltd. GED is an employee of ISCA Diagnostics Ltd.

## Notes

### Funding Statement

This study was funded by ISCA Diagnostics Limited.

### Author Declarations

Ethics approval was given by the Royal Devon University Healthcare NHS Foundation Trust, Exeter, Devon, United Kingdom. Human tissue samples were obtained for diagnostic purposes with informed written consent. Samples were tested, examined and disposed of, with informed written consent, and in accordance with the Human Tissue Act 2004, and the Human Tissue Authority Code of Practice. The tissue samples had been de-identified prior to use in our study.

## References

1. Thornton, C.R. The potential for rapid antigen testing for mucormycosis in the context of COVID-19. Expert Rev. Mol. Diagn. 2024, 3, 161–167.

2. Morrissey, C.O.; Kim, H.Y.; Granham, K.; Dao, A.; Chakrabarti, A.; Perfect, J.R.; Alastruey-Izquierdo, A.; Harrison, T.S.; Bongomin, F.; Galas, M.; et al. Mucorales: a systematic review to inform the World Health Organization priority list of fungal pathogens. Med. Mycol. 2024, 62, myad130.

3. WHO (2022). WHO fungal priority pathogens list to guide research, development and public health action (Geneva: World Health Organization). License: CC BY-NC-SA 3.0 IGO.

4. Casalini, G., Giacomelli, A., Antinori, S. The WHO fungal priority pathogens list: a crucial reappraisal to review the prioritisation. Lancet Microbe. 2024, 5, P717–724.

5. Cornely, O.A.; Alastruey-Izquierdo, A.; Arenz, D.; Chen, S.C.A.; Dannaoui, E.; Hochhegger, B.; Hoenigl, M.; Jensen, H.E.; Lagrou, K.; Lewis, R.E.; et al. Global guidelines for the diagnosis and management of mucormycosis: an initiative of the European Confederation of Medical Mycology in cooperation with the Mycoses Study Group Education and Research Consortium. Lancet Infect. Dis. 2019, 19, e405–e421.

6. Safiia, J., Díaz, M.C., Alshaker, H., Atallah, C.J., Sakr, P., Moshovitis, D.G., Nawio, A., Franceschi, A.E., Liakos, A., Koo, S. Recent advances in diagnostic approaches for mucormycosis. J. Fungi 2024, 10, 727.

7. Shen, H.; Cai, X.; Liu, J.; Yan, G.; Ye, Y.; Ding, R.; Wu, J.; Li, L.; Shen, Q.; Ma, Y.; et al. Case report: the clinical utility of metagenomic next-generation sequencing in mucormycosis diagnosis caused by fatal *Lichtheimia ramosa* infection in pediatric neuroblastoma. Front. Pediatr. 2023, 11, 1130775.

8. Shao, J.; Wan, Z.; Li, R.; Yu, J. Species identification and delineation of pathogenic *Mucorales* by Matrix-Assisted Laser Desorption Ionisation-Time of Flight Mass Spectrometry. J. Clin. Microbiol. 2018, 56, e01886–17.

9. Millon, L.; Herbrecht, R.; Grenouillet, F.; Morio, F.; Alanio, A.; Letscher-Bru, V.; Cassaing, S.; Chouaki, T.; Kauffman-Lacroix, C.; Poirier, P. et al. Early diagnosis and monitoring of mucormycosis by detection of circulating DNA in serum: retrospective analysis of 44 cases collected through the French Surveillence Network of Invasive Fungal Infections (RESSIF). Clin. Microbiol. Infect. 2016, 22, 810.

10. Vanbiervliet, Y., Aerts, R., Maessen, L., Wauters, J., Maertens, J., Lagrou, K. Laboratory innovations to diagnose invasive mould infections-what is relevant, what is not? Clin. Microbiol. Infect. 2026, 32, P715–P728.

11. Davies, G.E., Thornton, C.R. Development of a monoclonal antibody and a serodiagnostic lateral-flow device specific to *Rhizopus arrhizus* (Syn. *R. oryzae*), the principal global agent of mucormycosis in humans. J. Fungi 2022, 8, 756.

12. Thornton, C.R., Davies, G.E. Monoclonal antibodies can aid in the culture-based detection and differentiation of *Mucorales* fungi–the flesh-eating pathogens *Apophysomyces* and *Saksenaea* as an exemplar. Antibodies 2025, 14, 85.

13. Thornton, C.R., Davis, G.E., Dougherty, L. Development of a monoclonal antibody and a lateral-flow device for the rapid detection of a *Mucorales-*specific biomarker. Front. Cell. Infect. Microbiol. 2023, 13, 1305662.

14. Hudson, A.C., Corzo-Léon, D.E. Kalinina, I., Wilson, D., Thornton, C.R., Warris, A., Ballou, E.R. Characterization of the spatiotemporal localization of a pan-Mucorales-specific antigen during germination and immunohistochemistry. J. Infect. Dis. 2024, jiae375.

15. Rousselot, J.; Millon, L.; Scherer, E.; Bourgeois, N.; Imbert, S.; Dupont, D.; Debourgogne, A.; Maubon, D.; Bellanger, A.P.; Thornton, C.R. Detection of *Mucorales* antigen in bronchoalveolar lavage samples using a newly developed lateral-flow device. J. Clin. Microbiol. 2025, 63, e00226–25.

16. Thornton, C.R. Immunological Methods for Fungi. In Molecular and Cellular Biology of Filamentous Fungi: A Practical Approach, 1^st^ ed.; Talbot, N.J., Ed.; Oxford University Press: Oxford, UK, 2001; pp. 227–256.

17. Hudson, A.C., Rymer, B., Pradeep, S., Day, J.N., Corzo-León, D.E., Borman, A.M., Ballou, E.R., Thornton, C.R. Cutaneous mucormycosis confirmed using a *Mucorales*-specific monoclonal antibody: a case study. Front. Med. 2026, 1823222, in press. https://www.frontiersin.org/journals/medicine/articles/10.3389/fmed.2026.1823222/abstract

18. Thornton, C.R. Detection of the ‘big five’ mold killers of humans: *Aspergillus, Fusarium, Lomentospora, Scedosporium* and Mucormycetes. Adv. Appl. Microbiol. 2020, 110, 1–61.

19. Choudhary, H., Kaur, H., Singh, S., Singh, R., Muthu, V., Verma, R., Rudramurthy, S.M., Agarwal, R., Jain, S., Bal, A., et al. A novel indirect ELISA for serodiagnosis of mucormycosis using antigens from *Rhizopus arrhizus*. Mycoses 2024, 67, e13730.

20. Woodward, M.P., Young, W.W., Bloodgood, R.A. Detection of monoclonal antibodies specific for carbohydrate epitopes using periodate oxidation. J. Immunol. Methods 1985, 78, 143–153.

21. Alhajj, M., Zubair, M., Farhana, A. (2025) Enzyme Linked Immunosorbent Assay. [Updated 2023 Apr 23]. In: StatPearls [Internet]. Treasure Island (FL): StatPearls Publishing; 2025 Jan-. Available from: https://www.ncbi.nlm.nih.gov/books/NBK555922/

22. Dichtl, K., Seybold, U., Ormanns, S., Horns, H., Wagener, J. Evaluation of a novel *Aspergillus* antigen enzyme-linked immunosorbent assay. J. Clin. Microbiol. 2019, 57, e00136–19.

23. Egger, M., Penziner, S., Dichtl, K., Gornicec, M., Kriegl, L., Krause, R., Khong, E., Mehta, S., Vargas, M., Gianella, S., et al. Performance of the Euroimmun *Aspergillus* antigen ELISA for the diagnosis of invasive pulmonary aspergillosis in bronchoalveolar lavage fluid. J. Clin. Microbiol. 2022, 60, e0021522.

24. Skiada, A., Drogari-Apiranthitou, M., Roilides, E., Chander, J., Khostelidi, S., Klimko, N., Hamal, P., Chrenkova, V., Kanj., S.S., Zein, S.E., et al. A global analysis of cases of mucormycosis recorded in the European Confederation of Medical Mycology / International Society for Human and Animal Mycology (ECMM / ISHAM) Zygomyco.net registry. Mycopathologia 2025, 190, 53.

25. Erami, M., Mirhendi, H., Momen-Heravi, M., Sharif, A., Hezaveh, S.J.H., Matini, A.H., Ahsaniarani, A.H., Aboutalebian, S. Case report: COVID-19-associated mucormycosis co-infection with *Lomentospora prolificans*: the first case and review on multiple fungal co-infections during the COVID-19 pandemic. Front. Med. 2023, 10, 1078970.

26. Marques, D.S., Vaz, C.P., Branca, R., Campilho, F., Lamelas, C., Afonso, L.P., et al. *Rhizomucor* and *Scedosporium* infection post hematopoietic stem-cell transplant. Case Rep. Med. 2011, 2011, 830769.

27. Shand, J.M., Albrecht, R.M., Burnett, H.F., Miyake, A. Invasive fungal infection of the midfacial and orbital complex due to *Scedosporium apiospermum* and mucormycosis. J. Oral Maxillofac. Surg. 2004, 62, 231–234.

28. Song, Y., Zhou, M., Gong, Q., and Guo, J. *Scedosporium apiospermum* and *Lichtheimia corymbifera* co-infection due to inhalation of biogas in immunocompetent patients: a case series. Infect. Drug Resist. 2022, 15, 6423–6430.

29. Kanaujia, R., Muthu, V., Singh, S., Rudramurthy, S.M., Kaur, H. Rapidly progressive lung coinfection due to *Rhizopus* and *Scedosporium* in a diabetic marijuana smoker. Clin. Microbiol. Infect. 2023, 29, 51–53.

30. Marino, A., Calvo, M., Trovato, L., Scalia, G., Gussio, M., Consoli, U., Ceccarelli, M., Nunnari, G., Cacopardo, B. *Mucorales*/*Fusarium* mixed infection in hematologic patient with COVID-19 complications: an unfortunate combination. Pathogens 2023, 12, 304.

